# SARS-CoV-2 surveillance (09/2020 - 03/2021) in elementary schools and daycare facilities in Bavaria

**DOI:** 10.1101/2022.01.18.22269445

**Authors:** Anna Kern, Pia H. Kuhlmann, Stefan Matl, Markus Ege, Nicole Maison, Jana Eckert, Ulrich von Both, Uta Behrends, Melanie Anger, Michael C. Frühwald, Michael Gerstlauer, Joachim Woelfle, Antje Neubert, Michael Melter, Johannes Liese, David Goettler, Andreas Sing, Bernhard Liebl, Johannes Hübner, Christoph Klein, the COVID Kids Bavaria Consortium

## Abstract

Here we report our results of a multi-center, open cohort study (“COVID-Kids-Bavaria”) investigating the distribution of SARS-CoV-2 among children and staff in 99 daycare facilities and 48 elementary schools in Bavaria, Germany. Overall, 2568 children (1337 school children, 1231 preschool children) and 1288 adults (466 teachers, 822 daycare staff) consented to participate in the study and were randomly tested in three consecutive phases (September/October 2020, November/December 2020, March 2021). In total, 7062 throat swabs were analyzed for SARS-CoV-2 by RT-PCR. In phase I, only one daycare worker tested positive. In phase II, SARS-CoV-2 was detected in three daycare workers, two preschool children, and seven school children. In phase III, no sample tested positive. This corresponds to a positive test rate of 0.05% in phase I, 0.4% in phase II and 0% in phase III. After phase III, antibody testing was offered to 713 study participants in elementary schools. A seroprevalence rate of 7.7% (students) and 4.5% (teachers) was determined. We conclude that during the initial waves of the SARS-CoV-2 pandemic, the risk of a positive SARS-CoV-2 result correlated positively with the local 7-day incidence. Thus, an increased risk of SARS-CoV-2 transmission in the setting of daycare and elementary schooling was unlikely.

## Introduction

When the current coronavirus disease 19 (COVID-19) pandemic started spreading around the world, this scenario immediately invoked concerns with respect to earlier pandemic situations, such as the 1918 influenza pandemic. At that time, the influenza pandemic affected predominantly the younger age groups, and closing schools was an effective measure to control the spread of the disease [1]. Also, during seasonal influenza waves primarily young children played a significant role in disease spreading [2]. Initially, this raised concerns that children may be a major contributing factor for spreading severe acute respiratory syndrome coronavirus 2 (SARS-CoV-2).

Even though infection rates in children were eventually found to be as high as in adults [3], COVID-19 generally causes only mild disease in children, including infants [3, 4]. Many children remain completely asymptomatic [5]. Thus, the impact of asymptomatic individuals on operating schools and daycare facilities remained unclear. Many countries - including Germany - closed their schools leading to disrupted education for billions of learners worldwide [6].

Hence, the COVID Kids Bavaria study was initiated to investigate the occurrence and transmission rates of SARS-CoV-2 infections among children and staff in daycare facilities and elementary schools in Bavaria, Germany. Here, we report the occurrence of incident and prevalent cases in relation to the overall Bavarian incidence rates in three consecutive test phases between September 2020 and March 2021. We also determined the seroprevalence rate after completion of the PCR-based analysis.

## Methods

### Population

The COVID Kids Bavaria study is a large, multi-center, open cohort study investigating the distribution of SARS-CoV-2 among children and staff in daycare facilities and elementary schools in Bavaria, Germany. Six study centers were involved representing all Bavarian university children’s hospitals (Ludwig-Maximilians-Universität München, Technische Universität München, Augsburg, Erlangen, Regensburg, Würzburg).

Local ethics committees approved the study protocol and all related documents. COVID Kids Bavaria is registered with the German Clinical Trials Register (http://www.drks.de/DRKS00022380).

Bavaria is the largest of 16 federal states in Germany with a population of 13.2 million [7]. In 2408 elementary schools, approximately 442,000 children are being educated. The vast majority of these schools (2257) are public schools [8]. For representativeness, one public elementary school was selected from each of the 46 electoral districts, which divide the federal state of Bavaria in similar shares with respect to population size (Figure 1). Each elementary school was matched by two daycare facilities in the surrounding area (one nursery and one kindergarten or combined facilities). Facilities were characterized as urban or rural according to data from the Federal Statistical Bureau of Germany [9].

**Figure 1:**
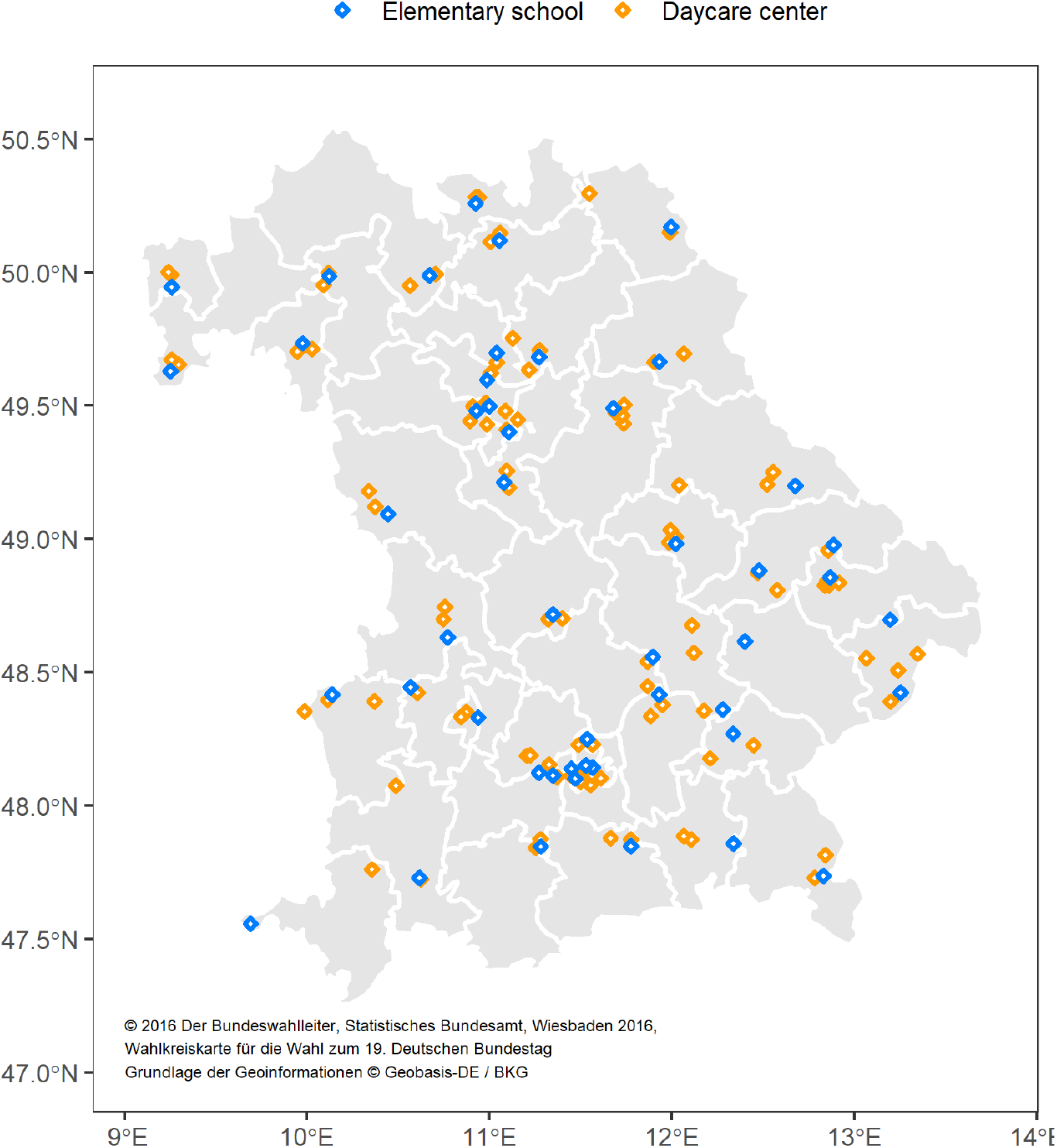
Participating facilities in Bavaria: The white lines delineate the 46 Bundestag constituencies with approximately equal population shares.

Participants were eligible for enrollment if they met the following inclusion criteria: child aged 1 to 10 years or teachers / daycare staff attending the participating facilities at the day of assessment and written informed consent provided by participants or their legal representatives. In addition, individuals had to be in a state of health permitting to visit the facilities as outlined by the latest corona guidelines from the department of health at the time of assessment [10].

Eligible participants were approached from June 2020 and recruitment was continued throughout all phases. Study visits occurred in each facility once in every phase. Phase I took place from 2020-09-23 till 2020-11-05, phase II from 2020-11-23 till 2020-12-16. Phase III was initially scheduled for 2021-01-25 – 2021-02-12 but was shifted to 2021-03-01 – 2021-- 03-26 due to lock down measures. Phase I was dominated by the SARS-CoV-2 wild type. Additionally, from December 2020 onwards variants of concern were detected in the German population (Alpha (B.1.1.7); Beta (B.1.351); Gamma (P.1)) until completion of throat swab sampling. To exclude contamination effects by infections acquired during holidays or lock down periods, each study phase started with a delay of at least two weeks from reopening of the facilities.

A pilot phase was conducted in July 2020 before the summer holiday break in two elementary schools to validate measurement instruments.

The majority of children enrolled in the pilot phase were visiting the last grade of elementary school. By design, the enrolled children of the last grade of the elementary school could not be followed up after the summer break. Therefore, the results of the pilot phase are reported separately.

For assessing non-response, an additional anonymous questionnaire was sent to parents of school children of 15 selected elementary schools via an online survey platform. Here, we asked for their reason to participate or not to participate in our study as well as compliance with SARS-CoV-2 protection measures and general demographics.

### PCR testing for SARS-CoV-2

Each visit entailed testing of a random sample for SARS-CoV-2 by oral throat swabs and subsequent pseudonymized RT-PCR analysis. Sample size for PCR-testing was calculated estimating a point prevalence of SARS-CoV-2 of 3% and a detection of every sixth case by testing only on a single day per class or group. This results in an expected proportion of positive tests of 0.5%. To determine the point prevalence on an alpha level of 5% with a power of 80%, 3000 samples per study phase were necessary.

For PCR testing, random subsamples were selected with 18 school children/preschool children and 4 teachers/daycare staff per facility during phase I. This figure was increased to 40 school children/20 preschool children and 20 teachers/10 daycare staff during the subsequent phases. In addition, the full set of pilot samples were subjected to PCR testing. PCR analyses for SARS-CoV-2 was conducted by laboratories of the Bavarian Health and Food Safety Authority (LGL) using the RT-PCR method. The tests detected at least 2 gene regions of the viral RNA. (E gene& S gene, RealStar SARS-CoV-2 RT-PCR Kit, Altona Diagnostics; E gene & Orf 1a gene, ampliCube Coronavirus SARS-CoV-2, Mikrogen Diagnostik; E gene & N2 gene, Xpert Xpress SARS-CoV-2,Cepheid; N gene & RdRp gene, Abbott RealTime SARS-CoV-2- Amplificaton Reagent Kit, Abbott Molecular). Positive test results were immediately reported to participants and local health authorities. Performance of Next Generation Sequencing was feasible for five samples.

Following a standardized questionnaire, secondary attack rates and infection circumstances were assessed retrospectively via telephone interviews with participants tested positive in the PCR.

### Seroprevalence substudy

To assess seroprevalence in our cohort, antibody-testing against SARS-CoV-2 was offered to staff and children in 15 selected elementary schools in June/July 2021.

Antibody testing was conducted by the LMU Klinikum, Department of Infectious Disease and Tropical Medicine, using a pan IgG-antibody test[11]. The N-Antigen was detected analysing capillary blood on standardized filter paper (Anti-SARS-CoV-2 N, Roche). At the same time participants completed a questionnaire regarding the history of a SARS-CoV-2 infection confirmed by laboratory testing.

### Statistical analyses

Descriptive statistics included summaries of participation rates at the facility and individual level. Key characteristics of participants (age, sex) were summarized as mean (range) or count (percentage). Missing values are listed in the respective tables; they were not imputed. Characteristics of responders and non-responders were compared by Fisher’s exact test. Odds ratios (OR) with 95%-confidence intervals (CI) were also calculated by Fisher’s exact test; Wilcoxon rank sum test was used to compare continuous variables. All statistical analyses were performed with R (version 4.0.0) [12].

## Results

Of all 149 enrolled facilities, 147 facilities were visited by study teams (Figure 1). The selected facilities represented urban (23 elementary schools, 54 daycare facilities) and rural areas (25 elementary schools, 45 daycare facilities). Two facilities were not visited as they were locked down during sampling periods.

Extrapolating from the size and number of facilities, we expected 10,723 school children, 682 teachers, 8586 preschool children, 1299 daycare staff to be eligible. Of these, 2568 (66.6%) children (1337 (34.7%) school children, 1231 (31.9%) preschool children) and 1288 (33.4%) adults (466 (12.1%) teachers, 822 (21.3%) daycare staff) consented to participate in the study and to provide throat swabs for SARS-CoV-2 testing (Figure2).

**Figure 2:**
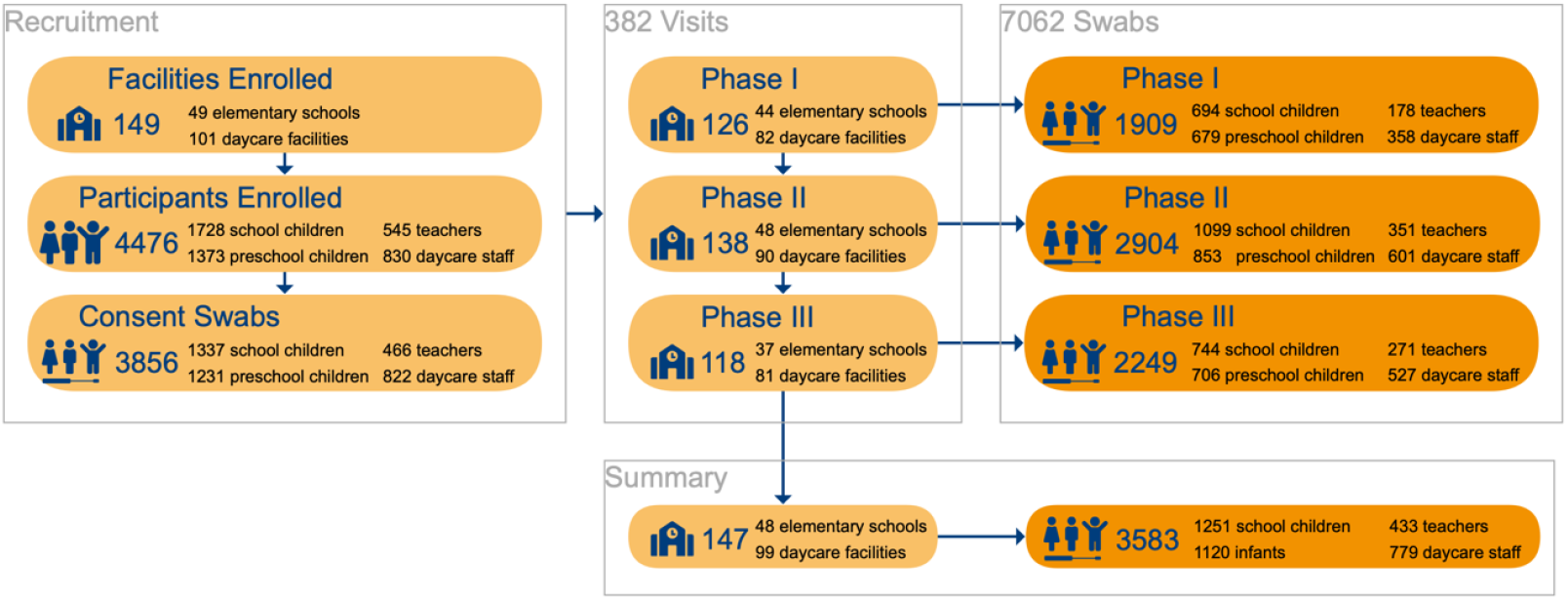
Participant Flow: Inclusion and participation of study individuals for throat swab-PCR-testing.

Preschool children covered an age range from 0 to 7 years and school children from 6 to 12 years (Table 1). Sex distribution was equal in children, whereas teachers and daycare staff were predominantly female (89% and 95%, respectively). The non-response assessment questionnaire was answered almost equally by the parents of participating and non-participating individuals (46.9% and 53.1% respectively). Participating and non-participating children were similar in demographics and their parents had a similar educational level and felt equally affected by the COVID-19 pandemic (Supplemental Table 1). Non-participation in the study was associated with considering general hygiene measures (e.g., wearing masks, social distancing, and washing hands) to be less important, lower interest in vaccinating their children against SARS-CoV-2 and being less anxious about contracting SARS-CoV-2. The most common reasons for parents not to participate were i) the wish to spare their children a throat swab and ii) rejection of their children to participate.

**Table 1:**
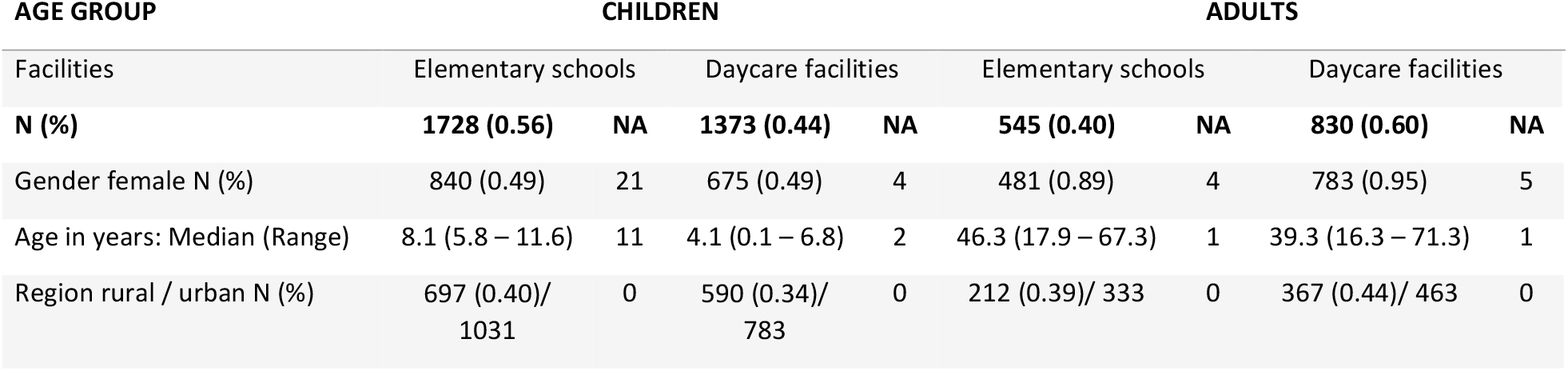
Population characteristics by study group: Number (N) and proportion (%) of participants; NA = not available (missing data)

| AGE GROUP | CHILDREN |  |  |  | ADULTS |  |  |  |
| --- | --- | --- | --- | --- | --- | --- | --- | --- |
| Facilities | Elementary schools |  | Daycare facilities |  | Elementary schools |  | Daycare facilities |  |
| **N (%)** | **1728 (0.56)** | **NA** | **1373 (0.44)** | **NA** | **545 (0.40)** | **NA** | **830 (0.60)** | **NA** |
| Gender female N (%) | 840 (0.49) | 21 | 675 (0.49) | 4 | 481 (0.89) | 4 | 783 (0.95) | 5 |
| Age in years: Median (Range) | 8.1 (5.8 – 11.6) | 11 | 4.1 (0.1 – 6.8) | 2 | 46.3 (17.9 – 67.3) | 1 | 39.3 (16.3 – 71.3) | 1 |
| Region rural / urban N (%) | 697 (0.40)/<br>1031 | 0 | 590 (0.34)/<br>783 | 0 | 212 (0.39)/ 333 | 0 | 367 (0.44)/ 463 | 0 |

PCR-testing for SARS-CoV-2 was performed in 7062 throat swab samples, 4775 (67.6%) samples from children (2537(35.9%) school children, 2238 (31.7%) preschool children) and 2287 (32,4%) samples from adults (800 (11.3%) teachers, 1487 (21.1%) daycare staff). Of these, 13 samples tested positive for SARS-CoV-2 with one daycare worker in phase I and three daycare workers, two preschool children, and seven school children in phase II (Figure 3). The throat swab samples obtained in phase III yielded no positive SARS-CoV-2 PCR results. These figures correspond to a positive test rate of 0.05% in phase I, 0.4% in phase II and 0% in phase III.

**Figure 3:**
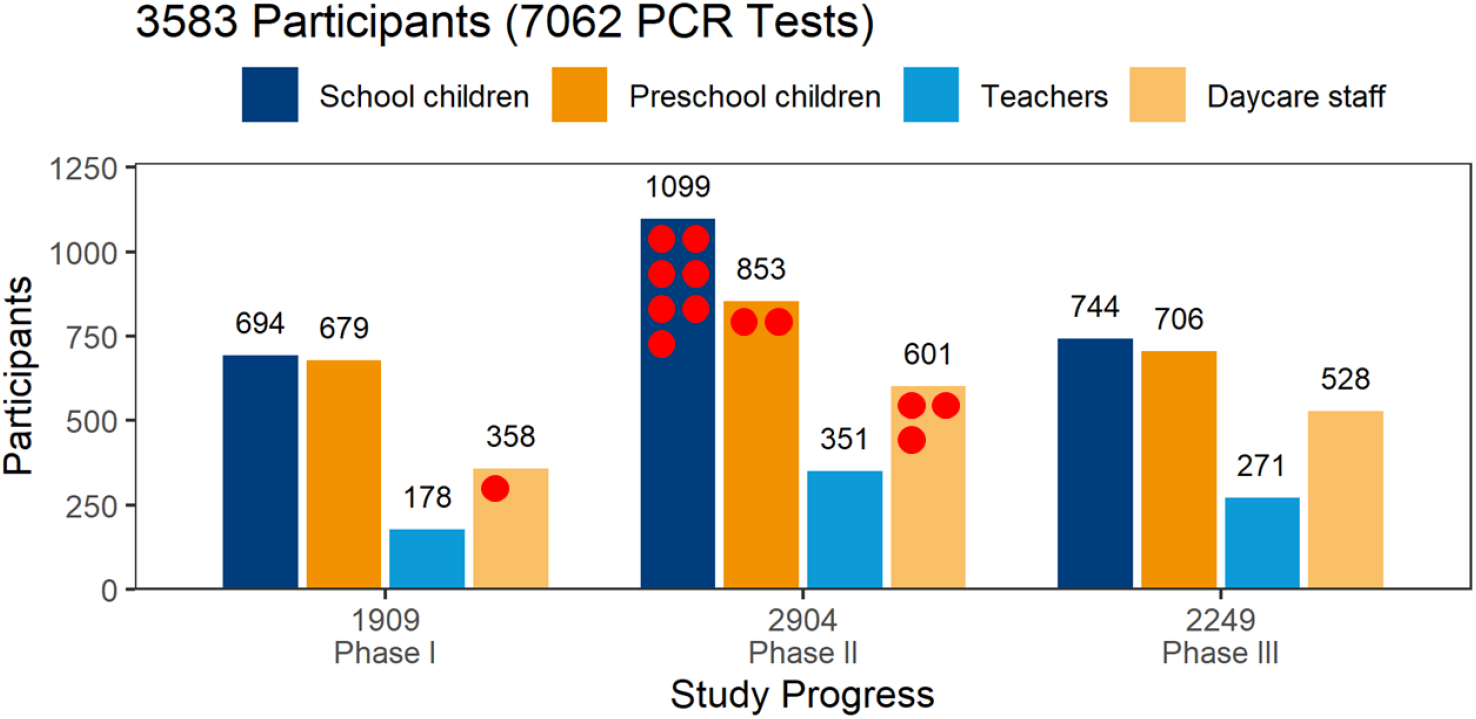
Participants tested by phase and study group: Numbers of participants tested are given by study group and study phase. Altogether 7062 tests were performed with no available result in 9 tests. The 13 positive PCR tests are marked by red dots.

Four daycare workers and six children were newly detected by our study. Three other children with positive PCR tests turned out to have been diagnosed with SARS-CoV-2 by other measures in the past. They were allowed to reenter their school after completing a 14-days isolation period and still tested positive after readmission (Table 2).

**Table 2:** Characteristics of participants with positive test result: § The test was performed after the individual had been tested positive previously and had been isolated for 14 days. * Virus variants were not determinable in most cases due to low number of viral copies in the sample (Ct values mostly <30).

| PHASE | FACILITY | AGE<br>GROUP | CT-VALUE<br><30 | VIRUS<br>VARIANT | SUPPOSED MODE<br>OF TRANSMISSION | INITIAL<br>SYMPTOMS | SUBSEQUENT<br>SYMPTOMS | SUPPOSED<br>FURTHER<br>TRANSMISSION | DETAILS |
| --- | --- | --- | --- | --- | --- | --- | --- | --- | --- |
| I | Daycare | Adult | No | Unknown * | Colleague at work | Yes | Yes | No |  |
| II | Daycare | Adult | No | Unknown * | Colleague/child at work | No | Yes | Family |  |
| II | Daycare | Adult | No | Unknown * | Colleague/child at work | No | Yes | No |  |
| II | Daycare | Adult | No | Unknown * | Colleague/child at work | No | No | No |  |
| II | Daycare | Child | No | Unknown * | Family | No | No | No | Prior infection<sup>§</sup> |
| II | Daycare | Child | No | B.1.177 | Family | No | Yes | No |  |
| II | School | Child | No | Unknown * | Family | No | No | No | Prior infection<sup>§</sup> |
| II | School | Child | No | Unknown * | Family | No | Yes | No |  |
| II | School | Child | yes | B1.160 | Family | No | Yes | Family | In quarantine<br>when test result<br>arrived |
| II | School | Child | No | Unknown * | Not known | No | No | No |  |
| II | School | Child | No | Unknown * | Not known | No | Not known | Not known | Prior infection<sup>§</sup> |
| II | School | Child | No | B1.160 | Not known | No | No | No | In quarantine<br>shortly before<br>testing |
| II | School | Child | No | Unknown * | Not known | Yes | No | No |  |

Two potential clusters with two positively tested children each within one school, but different classes, were identified. In the retrospective case analysis, five families stated that their children were presumably infected by other family members. With a cycle threshold (ct) of <30, only one child was presumably highly infectious at the time of testing. This child had already been quarantined as a contact person on the subsequent day before our test result arrived (Table 2).

The results of the pilot study were analyzed separately (see Methods). In July 2020, two elementary schools were visited, and 60 swabs were taken in 32 school children and 28 teachers. At the time, schools reduced the number of students per class following public health policy measures. Therefore, classes were divided into two groups. In one group of fourth-grade students, two children tested positive, whereas the other group could not be tested as they had been quarantined shortly before.

The countrywide incidence of COVID-19 varied considerably over the study phases (Figure 4). The local 7-day incidence used for calculation refers to the 7-day incidence of the administrative district of each individual sampling site on the day of testing.

**Figure 4:**
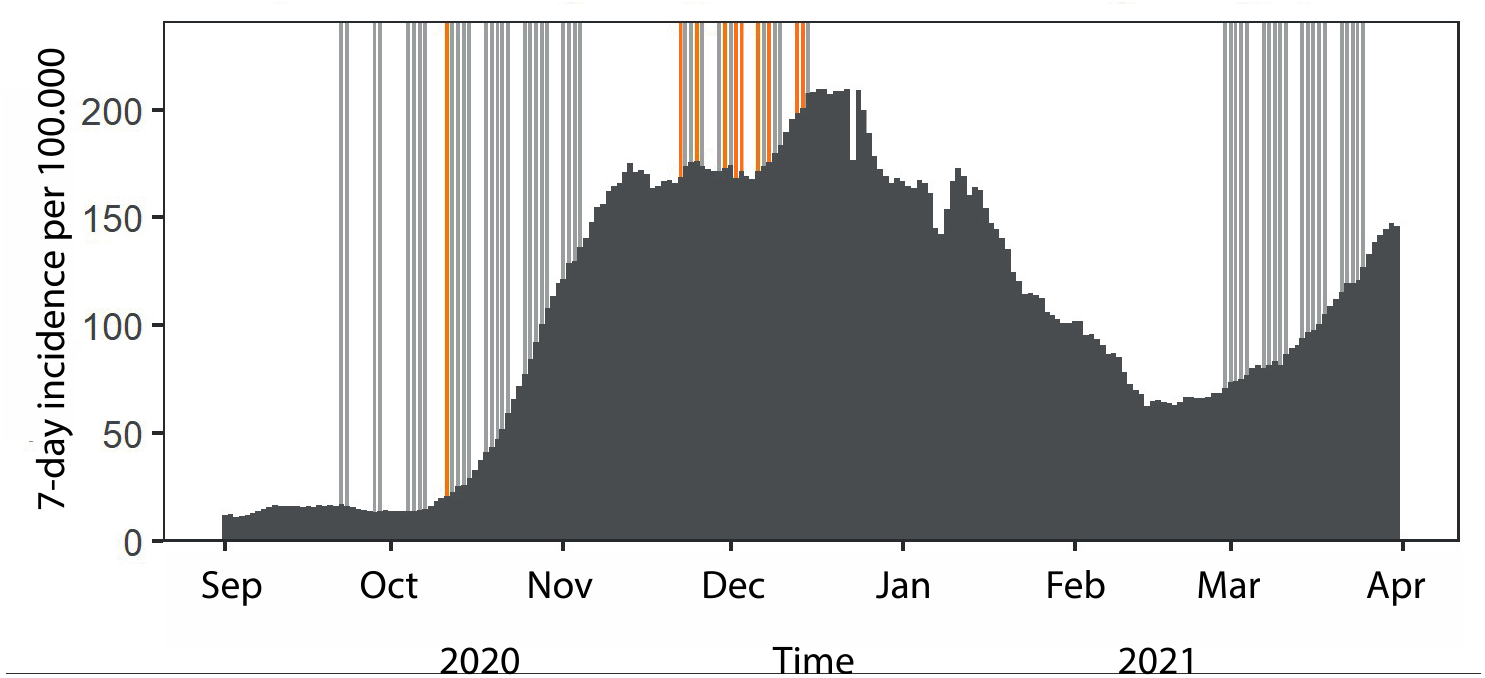
Timeline of testing: Dark grey bars denote the daily 7-day incidence/100,000 in Bavaria. The vertical lines reflect the days of PCR testing in Phase I, II and III, respectively. Days with positive PCR test results for SARS-CoV-2 are marked by orange lines.

During phase I the mean local 7-day-incidence was 55/100,000 (range: 4 – 255/100.000), during phase II 184/100,000 (range: 84-580/100.000) and during phase III 77/100,000 (range: 12-238/100.000), yielding a mean local 7-day-incidence of 115/100,000 over all tests performed. Positive PCR tests were obtained on days with a local 7-day incidence of 212/100,000 on average, whereas negative PCR tests corresponded to an average 7-day incidence of 115/100,000 (Figure 5). A positive PCR test result was strongly associated with a local 7-day incidence of more than 100/100,000 as compared to less than 100/100,000 (OR=10.3 [1.5 - 438], p<0.005).

**Figure 5:**
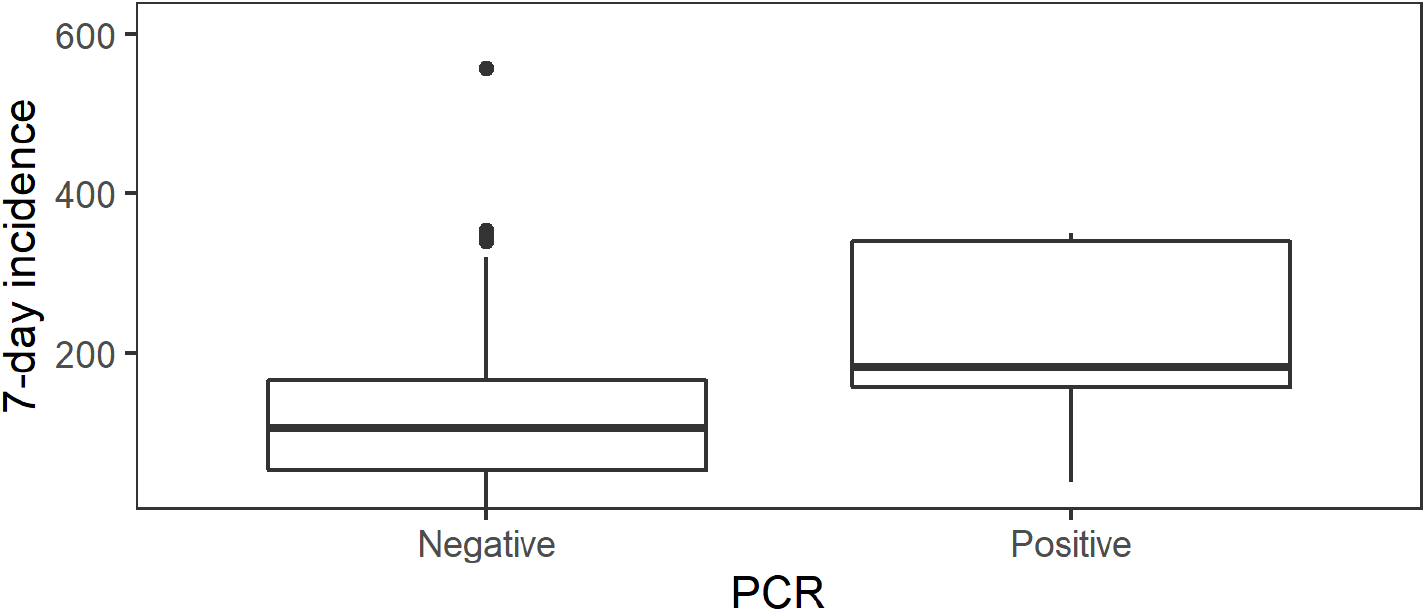
PCR test results in the context of local 7-day incidence numbers: The local 7-day incidence per 100,000 individuals differs between days when negative and positive PCR tests results were obtained in our study (p<0.0005).

Upon completion of the three phases of PCR-testing, IgG antibody-testing for SARS-CoV-2 was performed in 713 individuals attending elementary schools (511 (71.7%) children, 202 (28.3%) teachers).Of these, 39 school children and 9 teachers tested positive for SARS-CoV-2- antibodies, whereas negative results were obtained in 470 school children and 193 teachers. This corresponds to a seroprevalence of 7.7% in school children and 4.5% in teachers. In 2 samples from school children, serum antibody assays failed for technical reasons and therefore measurements were not available.

Of all individuals tested for SARS-CoV-2-antibodies, 438 provided information on whether a potential previous infection confirmed by laboratory testing has occurred or not (Table 3). The recall of an infection with SARS-CoV-2 yielded a false negative rate of 50% in adults and 56% in children.

**Table 3:** Recall of SARS-CoV-2 infection and measured antibodies against SARS-CoV-2

|  | Detection of |  | Recall of positive PCR test |  | False negative | False positive |
| --- | --- | --- | --- | --- | --- | --- |
| Age group | SARS-CoV-2 antibodies |  | Yes | No |  |  |
| Adults | Yes | 6 | 3 | 3 | 50% |  |
|  | No | 117 | 3 | 114 |  | 3% |
| Children | Yes | 27 | 12 | 15 | 56% |  |
|  | No | 288 | 2 | 286 |  | 1% |

## Discussion

The COVID Kids Bavaria study assessed the occurrence of SARS-CoV-2 in healthy individuals attending elementary schools and day care facilities in three different phases covering the second and third wave of the COVID-19 pandemic in Bavaria. Of 7,062 PCR-tests in 3,856 participants, 13 yielded a positive test result (4 daycare workers, 2 preschool children, 7 school children), of whom three children were known cases testing still positive after a required isolation period of 14 days. A positive PCR test result was strongly associated with a local 7-day incidence of more than 100/100,000 as compared to less than 100/100,000 (OR=10.3 [1.5 - 438], p<0.005). Half of the individuals with detectable IgG antibodies against SARS-CoV-2 were unaware of a previous infection.

Essentially, we did not intend to quantify the overall prevalence of disease within the specified target population, as this is already done by other scientific studies [13, 14] and the health authorities [15]. Rather we aimed to assess the spread of the disease in healthy individuals attending day care facilities and elementary schools on a regular basis. A strength of our study is the broad coverage of urban and rural areas and the equal representation of all Bavarian districts. This has been facilitated by a collaborative effort of all University Children’s Hospitals of Bavaria, the support of the health authorities, and the involved facilities. An anonymous non-responder questionnaire showed no significant differences with respect to demographics and experience of personal limitations due to restrictions of everyday life. However, participants and non-participants differed in their perception of their personal risk and of the necessity of hygiene measures. These moderate differences were expected and indicate a minor but no major selection bias.

The initial sample size calculation was based on the assumption that 0.5% of PCR samples would be positive. This figure was derived from an estimated point prevalence of 3% [16] and an average incubation period of 6 days [17]. When testing an individual only on one day, as in our study by design, 5 out of 6 individuals might escape. Retrospectively the assumed figure of positive samples was an overestimation, and the projected sample size was not reached in phase I due to a low recall rate and in phase III due to lockdown measures. However, intensified recruitment in phase II led to an inclusion of 2900 individuals and a detection of 0.4% positive samples thereby almost meeting the prior assumptions.

On the other hand, the low number of detected cases is informative in itself as it suggests that established hygiene measures worked reliably and prevented daycare facilities and elementary schools from major outbreaks. These findings build upon the results of a previous study conducted in the Munich metropolitan area [18]. Moreover, the risk of asymptomatic SARS-CoV-2 infections in children visiting daycare or elementary schools was rather low, as hardly any new cases were detected. Only nine children were tested positive, three of these were known cases detected upon recovery and after 14 days of isolation. These three children had a low virus load as indicated by Ct values > 30 and thus were unlikely to spread SARS-CoV-2.

Our detection rate was closely related to the local incidence numbers in the respective administrative districts of Bavaria, which is illustrated by the particularly strong association of detection of cases in our population with the concurrent local incidence values above 100/100,000. Together with the exact timing of the detection of positive samples (Figure 4), this demonstrates that the COVID Kids Bavaria study mirrored well the spread of SARS-CoV-2 across Bavaria. Similar findings were reported by a study from Public Health England, which describes a strong association of SARS-CoV-2 outbreaks among staff and students in educational settings during June / July 2020 in England with the regional COVID-19 incidence [19]. Studies from other countries have also highlighted that community transmission of SARS-CoV-2 is a risk factor for transmission in daycare facilities and schools [3, 20]. Collectively, these results are in line with the concept that the spread of SARS-CoV-2 in the population is not primarily “driven” by children attending elementary schools and daycare facilities in Germany [18, 21-23].

The study was not designed to track chains of infection, to assess local outbreaks, or to determine the secondary attack rate. Our random sampling strategy thus cannot offer a complete picture of the epidemic activity in entire school classes and daycare groups. Nevertheless, retrospective interviews with parents suggested that transmission of the virus to children occurred predominantly at home and not within daycare facilities or elementary schools. Secondary attacks within facilities were not reported, indicating that daycare facilities and schools did not pose a higher risk of SARS-CoV-2 transmissions during the second and third wave of the pandemic. This is in line with findings of a case control study from the U.S. in children and adolescents: A positive SARS-CoV-2 test was not associated with in-person school or childcare attendance during the preceding two weeks [24].

Furthermore, a recent report of the European Centre for Disease Prevention and Control (ECDC) showed that SARS-CoV-2 outbreaks within schools led to clusters of usually less than 10 persons and was mainly observed in secondary and not in primary schools [3]. In our survey, only a single individual was found to have a high viral load (i.e., a Ct value below 30) and thus a marked potential to spread the virus. Of note, even before the return of our PCR test result, this child had been quarantined because a close SARS-CoV-2 positive contact person had been identified by the health authorities. In other words, only one person day of the about 7,000 person days covered by our tests bore a marked risk of spreading the disease.

Our study population was characterized by the absence of symptoms suggestive of a SARS-CoV-2 infection as we deliberately tested children and adults who were allowed to attend their respective facilities only in good health. This is in sharp contrast to the predominant testing strategy of the health authorities, which focused mainly on symptomatic individuals or those with a high likelihood of relevant exposure. Therefore, asymptomatic individuals might have been missed by the authorities, thereby underestimating the true incidence in the entire population systematically. Conversely, our approach missed symptomatic individuals as these were not present at their facilities during the assessment.

Upon completion of the PCR-test phases, we added a serological study module to estimate the numbers of recorded and unrecorded cases in the population of school children and their teachers. The detected seroprevalence of 7.7% among school children in summer 2021 corresponds to the figure of 8.4% determined in 15,771 children in a seroepidemiological study in Bavaria, Germany [25] and 7.8% determined in 2500 Swiss children [26] in fall 2020. The proportion of unrecorded cases, i.e., individuals unaware of a previous infection, was about 50%. Thus, we might have missed every second case of a SARS-CoV-2 infection in our population. This could be explained by our focus on asymptomatic individuals and the short time window of assessment, i.e., just one day per individual. Nevertheless, the share of unrecorded cases was much lower as compared to an earlier seroepidemiological study in Bavaria: In fall 2020, infections among children up to 18 years old were 6-fold higher than the actual reported incidence. In the beginning of 2021, the numbers were still three to four times higher [25, 27]. This gradual decline may reflect an increase in the coverage of tests, particularly in asymptomatic individuals. This notion is supported by our antibody sub-study with an even lower share of unrecorded cases.

At the time of writing this manuscript, Health Authorities in Bavaria have intensified the screening efforts by rolling out RT-PCR pool testing and Antigen testing 2-3 times per week, thereby missing hardly any incident case in children, including asymptomatic individuals. In week 48 (December) 2021, the corresponding 7-day-incidence values were 1,172 in school children (aged 6 – 11 years) and 579 in middle-aged adults (35-59 years) [28]. Consequently, our data from the first waves of the COVID pandemic cannot easily be compared to the current situation. Moreover, at the time of our field phases few adults and virtually no children were immunized, and infection dynamics have been changing substantially with newly emerging SARS-CoV-2 strains characterized by increased transmissibility.

Nevertheless, the study is in line with the idea that hygiene measures and testing strategies during the early waves of Covid contained the pandemic in elementary schools and daycare settings, particularly when the Bavarian incidence was high (i.e., phase II during the second wave). With systematic and comprehensive testing strategies [29, 30], maintaining hygiene measures and increasing vaccination rates we may be in a better situation to face the currently prevailing Delta and Omicron variants without falling back to less sophisticated measures such as extensive and complete closing of schools and daycare facilities. While closing schools may have been a rational strategy during the influenza pandemic about 100 years ago, today, we have access to highly improved diagnostic and preventive measures. Our data, along with other studies, provide an argument for reserving complete shutdown of schools and daycare facilities as an ultima ratio measure.

## Data Availability

All data produced in the present study are available upon reasonable request to the authors.

## Acknowledgements

We thank all study participants for accepting our invitation and being part of this assessment. We acknowledge great financial support by the Bavarian Ministry of Science and the Arts. The study has also been supported by the Care-for-Rare Foundation and its philanthropic partners. We are indebted to our enthusiastic clinical and laboratory teams, without their help the study would not have been possible.

## References

1. Short, K.R., K. Kedzierska, and C.E. van de Sandt, Back to the Future: Lessons Learned From the 1918 Influenza Pandemic. Front Cell Infect Microbiol, 2018. 8: p. 343.

2. Tsang, T.K., et al., Individual Correlates of Infectivity of Influenza A Virus Infections in Households. PLoS One, 2016. 11(5): p. e0154418.

3. ECDC, COVID-19 in children and the role of school settings in transmission - second update. 2021: Stockholm.

4. Götzinger, F., et al., COVID-19 in children and adolescents in Europe: a multinational, multicentre cohort study. The Lancet Child & Adolescent Health, 2020. 4(9): p. 653–661.

5. Dong, Y., et al., Epidemiology of COVID-19 Among Children in China. Pediatrics, 2020. 145(6).

6. UNESCO, School closures caused by Coronavirus (Covid-19). 2020, Unesco.

7. Statistisches Bundesamt (Destatis). 14. koordinierte Bevölkerungsvorausberechnung nach Bundesländern - Moderate Entwicklung der Geburtenhäufigkeit, der Lebenserwartung und des Wanderungssaldos (G2-L2-W2). 2019 [cited 2021 23rd of December]; Available from: https://service.destatis.de/laenderpyramiden/.

8. Bayerisches Staatsministerium für Unterricht und Kultus, I 1. Überblick. Bayerns Schulen in Zahlen 2018/2019, 2019. Heft 67(Schriften des Bayerischen Staatsministeriums für Unterricht und Kultus, Reihe A, Bildungsstatistik,): p. 6.

9. Thünen-Institut für Ländliche Räume, Landatlas (http://www.landatlas.de). 2021: Braunschweig.

10. Bayerisches-Staatsministerium für Unterricht und Kultus. Coronavirus aktuell-Aktualisierter Rahmen-Hygieneplan für bayerische Schulen. 2021 [cited 2021 14th of December]; Available from: https://www.km.bayern.de/allgemein/meldung/7061/aktualisierter-rahmen-hygieneplan-fuer-bayerische-schulen.html.

11. Beyerl, J., et al., A dried blood spot protocol for high throughput analysis of SARS-CoV-2 serology based on the Roche Elecsys anti-N assay. EBioMedicine, 2021. 70: p. 103502.

12. R Core Team, R: A language and environment for statistical computing. 2020, R Foundation for Statistical Computing: Vienna, Austria.

13. Tonshoff, B., et al., Prevalence of SARS-CoV-2 Infection in Children and Their Parents in Southwest Germany. JAMA Pediatr, 2021. 175(6): p. 586–593.

14. Gudbjartsson, D.F., et al., Spread of SARS-CoV-2 in the Icelandic Population. N Engl J Med, 2020. 382(24): p. 2302–2315.

15. Robert Koch-Institut; Abteilung für Epidemiologie und Gesundheitsmonitoring (2021). COVID-19: Fallzahlen in Deutschland und weltweit -Fallzahlen in Deutschland -Stand: Dienstag 14.12.2021, 00:00 Uhr (online aktualisiert um 06:30 Uhr). 2021 [cited 2021 14th of December]; Available from: https://www.rki.de/DE/Content/InfAZ/N/Neuartiges_Coronavirus/Fallzahlen.html.

16. Pollán, M., et al., Prevalence of SARS-CoV-2 in Spain (ENE-COVID): a nationwide, population-based seroepidemiological study. The Lancet, 2020. 396(10250): p. 535–544.

17. Lauer, S.A., et al., The Incubation Period of Coronavirus Disease 2019 (COVID-19) From Publicly Reported Confirmed Cases: Estimation and Application. Ann Intern Med, 2020. 172(9): p. 577–582.

18. Hoch, M., et al., Weekly SARS-CoV-2 Sentinel Surveillance in Primary Schools, Kindergartens, and Nurseries, Germany, June-November 2020. Emerg Infect Dis, 2021. 27(8): p. 2192–2196.

19. Ismail, S.A., et al., SARS-CoV-2 infection and transmission in educational settings: a prospective, cross-sectional analysis of infection clusters and outbreaks in England. The Lancet Infectious Diseases, 2021. 21(3): p. 344–353.

20. Macartney, K., et al., Transmission of SARS-CoV-2 in Australian educational settings: a prospective cohort study. The Lancet Child & Adolescent Health, 2020. 4(11): p. 807–816.

21. Haag, L., et al., Prevalence and Transmission of Severe Acute Respiratory Syndrome Coronavirus Type 2 in Childcare Facilities: A Longitudinal Study. J Pediatr, 2021. 237: p. 136–142.

22. Buchholz, U., et al., Epidemiologie von COVID-19 im Schulsetting. 2021(13): p. 3--16.

23. Kern, A., et al., Welche Rolle spielen Kinder in Schulen und Kindertagesstätten bei der Übertragung von SARS-CoV-2? – Eine evidenzbasierte Perspektive. Bundesgesundheitsblatt - Gesundheitsforschung - Gesundheitsschutz, 2021. 64(12): p. 1492–1499.

24. Hobbs, C.V., et al., Factors Associated with Positive SARS-CoV-2 Test Results in Outpatient Health Facilities and Emergency Departments Among Children and Adolescents Aged< 18 Years—Mississippi, September–November 2020. 2020. 69(50): p. 1925.

25. Hippich, M., et al., A public health antibody screening indicates a marked increase of SARS-CoV-2 exposure rate in children during the second wave. Med (N Y), 2021. 2(5): p. 571–572.

26. Ulyte, A., et al., Clustering and longitudinal change in SARS-CoV-2 seroprevalence in school children in the canton of Zurich, Switzerland: prospective cohort study of 55 schools. BMJ, 2021. 372: p. n616.

27. Hippich, M., et al., A Public Health Antibody Screening Indicates a 6-Fold Higher SARS-CoV-2 Exposure Rate than Reported Cases in Children. Med (N Y), 2021. 2(2): p. 149–163 e4.

28. Bayerisches Landesamt für Gesundheit und Lebensmittelsicherheit 2020. Corona-Fälle in Bayern - 6. Inzidenz nach Alters-Gruppen,. 2021 14th of December, 2021 [cited 2021 14th of December].

29. Joachim, A., et al., Pooled RT-qPCR testing for SARS-CoV-2 surveillance in schools - a cluster randomised trial. EClinicalMedicine, 2021. 39: p. 101082.

30. Forster, J., et al., Feasibility of SARS-CoV-2 Surveillance Testing Among Children and Childcare Workers at German Day Care Centers: A Nonrandomized Controlled Trial. JAMA Network Open, 2022. 5(1): p. e2142057–e2142057.

