## Supplemental Table 1 for "SARS-CoV-2 surveillance (09/2020 - 03/2021) in elementary schools and daycare facilities in Bavaria"

### Supplementary Data

Supplemental Table 1: Results of non-response assessment questionnaire

| **Dichotomized Answers were applicable** | **Question** | **p** | **OR** | **Participants** | | **Non-Participants** | |
| --- | --- | --- | --- | --- | --- | --- | --- |
| rather yes / rather no | feeling affected by the pandemic | 0,910 | 0,96 | 214 | 45 | 244 | 49 |
| yes / no | personal freedom | 0,814 | 0,94 | 218 | 41 | 249 | 44 |
| yes / no | childcare | 0,169 | 1,44 | 230 | 29 | 248 | 45 |
| yes / no | professional life | 0,147 | 1,30 | 137 | 122 | 136 | 157 |
| yes / no | family life | 0,002 | 1,75 | 174 | 85 | 158 | 135 |
| yes / no | health | 0,288 | 0,82 | 88 | 171 | 113 | 180 |
| rather yes / rather no | agreement to general mitigation strategies | <0,001 | 2,93 | 200 | 59 | 157 | 136 |
| rather necessary / rather not necessary | distancing rules | <0,001 | 12,67 | 254 | 3 | 253 | 38 |
| rather necessary / rather not necessary | hand hygiene | 0,003 | 6,89 | 255 | 2 | 277 | 15 |
| rather necessary / rather not necessary | wearing a face mask | <0,001 | 6,14 | 240 | 17 | 204 | 89 |
| rather necessary / rather not necessary | frequent air ventilation in closed rooms | <0,001 | 51꙳ | 257 | 0 | 267 | 25 |
| rather necessary / rather not necessary | using the Corona Warn App | <0,001 | 2,55 | 117 | 140 | 72 | 220 |
| rather yes / rather no | fear of SARS-CoV-2 infection | <0,001 | 2,27 | 143 | 116 | 103 | 190 |
| rather yes / rather no | vaccinate child against SARS-CoV-2 | <0,001 | 3,00 | 179 | 80 | 125 | 168 |
| <= 40 y / > 40 y | age | <0,001 | 0,41 | 88 | 170 | 157 | 123 |
| yes / no | birthplace Germany | 0,017 | 1,89 | 234 | 25 | 243 | 49 |
| <= 12 y / >= 13 y | education (years) | 0,270 | 0,79 | 56 | 202 | 75 | 213 |
| ꙳ “Haldane-Anscombe” (HA) correction [1,2] | | | | | | | |

1. Haldane, J.B., *The estimation and significance of the logarithm of a ratio of frequencies.* Ann Hum Genet, 1956. **20**(4): p. 309-11.

2. Anscombe, F.J., *On Estimating Binomial Response Relations.* Biometrika, 1956. **43**(3/4).
